# Efficacy and Safety of Albendazole 400 mg for 30 Days in Adults Patients with Low *Loa loa* Microfilaremia: A Non-Inferiority Randomized Controlled Trial Compared to Ivermectin

**DOI:** 10.1101/2024.08.16.24310835

**Authors:** Luccheri Ndong Akomezoghe, Noé Patrick M’Bondoukwé, Denise Patricia Mawili Mboumba, Jacques Mari Ndong Ngomo, Moutombi Ditombi Bridy Chesli, Hadry Roger Sibi Matotou, Valentin Migueba, Marielle Karine Bouyou Akotet

**Affiliations:** Department of Parasitology-Mycology-Tropical Medicine, Faculty of Medicine, Université des Sciences de la Santé, BP 4009, Libreville - Gabon; Unité Mixte de Recherche sur les Agents Infectieux et leur Pathologie (UMRAIP), Université des Sciences de la Santé, Owendo, Gabon; Centre de REcherche biomédicale en pathogènes Infectieux et Pathologies Associées (CREIPA), Libreville, Gabon

## Abstract

**Background:** *Loa loa* infection is endemic in central African countries like Gabon and in West Africa. Treatment typically involves the use of ivermectin and albendazole, with albendazole often used to reduce microfilaremia in individuals with high microfilaremia before administering ivermectin. This study aims to evaluate the efficacy and safety of albendazole in patients with low microfilaremia.

**Methodology and principal findings:** The study was conducted from November 7 to April 1 across 31 villages in the Woleu-Ntem province of northern Gabon. Following a questionnaire, venous blood was collected in EDTA tubes for *Loa loa* diagnosis. Eligible individuals were randomized into two treatment groups and followed for 30 days. One group received daily albendazole tablets (400 mg), while the other received a single dose of ivermectin (200μg/kg). The study reported a 33.0% prevalence of *Loa loa* infection in northern Gabon. In the per-protocol analysis, the mean microfilaremia decreased significantly by 82.3% and 90.4% in the ALB and IVM groups, respectively (p˂ 0.001). The risk difference between the treatments was 8.1% [95% CI: 16.8; −0.6%]. For the intention-to-treat analysis, the mean microfilaremia decreased significantly by 82.4% and 90.8% in the ALB and IVM groups, respectively (p˂ 0.001), with a risk difference of 8.4% [95% CI: 16.2; 0.6%]. Eosinophil rates decreased by day 30 in both groups, though not significantly different (p>0.05).

**Conclusions/Significance:** The prevalence of *Loa loa* infection (33.05%) is notably high in northern Gabon. Albendazole demonstrated microfilaricidal activity in individuals with low *Loa loa* microfilaremia. However, its efficacy appears inferior to that of ivermectin and seems to diminish at very low microfilarial loads.

**Author summary:** *Loa loa* infection is endemic in central African countries, including Gabon, and across West Africa. Despite the absence of specific treatments developed for *Loa loa*, current therapeutic approaches predominantly rely on diethylcarbamazine (DEC), ivermectin (IVM), and albendazole (ALB). Although ALB is more readily available, it is typically reserved for patients with high microfilaremia due to potential severe adverse effects following treatment with IVM. This study aimed to evaluate the efficacy and safety of ALB as an alternative treatment for patients with low microfilaremia compared to IVM. The findings suggest that ALB could serve as a viable alternative for the treatment of microfilaremic loiasis. Moreover, ALB treatment demonstrated adequate clinical efficacy and safety comparable to that of IVM.

## Introduction

Loiasis is a neglected filarial parasitic disease endemic to West and Central Africa [1]. It is caused by the *Loa loa* worm, with adult worms residing in the skin or intermuscular fascia and microfilariae found in the blood. Over 10 million people are estimated to be carriers of this parasite [2]. Common clinical manifestations of loiasis include Calabar swelling, eyeworm, subconjunctival migration, subcutaneous crawling, and pruritus [3,4]. There are no specific drugs developed exclusively for *Loa loa*; however, current treatment strategies primarily rely on the use of diethylcarbamazine (DEC), ivermectin (IVM), and albendazole (ALB).

The World Health Organization (WHO) has implemented the Community-Directed Treatment with Ivermectin (CDTI) strategy to eradicate onchocerciasis and lymphatic filariasis [5,6]. However, in regions where onchocerciasis coexists with loiasis, CDTI poses a risk to individuals with *Loa loa* hypermicrofilaremia, defined as a parasite density higher that 8,000 microfilariae per milliliter of blood (mf/mL). Patients with this condition may experience severe and/or serious adverse events (SAEs) following IVM treatment, making areas with onchocerciasis-loiasis co-endemicity ineligible for CDTI [7,8]. In areas where onchocerciasis is hypoendemic, such as Gabon, a Test and Treat strategy is considered an alternative [9–11]. Several studies have been conducted to evaluate treatments that could safely reduce high *Loa loa* microfilaremia, thereby making populations eligible for IVM treatment. Different albendazole (ALB) regimens were tested using various designs. In Cameroon, administering ALB 400 mg for three days showed a reduction in microfilaremia at day 90 [12]. When patients with high *Loa loa* microfilaremia were given two or six doses of ALB 800 mg every two months, a decrease in microfilaremia was observed in patients receiving six doses after four months [13]. Recently in Gabon, a five-week regimen of ALB 400 mg showed a reduction in microfilaremia comparable to a treatment with ALB 400 mg for three weeks coupled with IVM [14]. Since the conclusion of the onchocerciasis control program activities in Gabon, IVM is not readily accessible to physicians and their patients. On one hand, these studies highlight the microfilaricidal effect of ALB; on the other hand, they underscore the lack of a standardized protocol for treating microfilaremic loiasis.

A significant body of research suggests that loiasis should be regarded as a major health issue [15,16]. Most loiasis patients have low microfilaremia (< 8000 mf/mL) and reside in rural areas [17,18]. In these regions, rural communities primarily access ALB through local dispensaries, which are their main healthcare facilities. The local populations primarily engage in agriculture, fishing and hunting, which generally do not provide substantial income. Clinically, low or absent microfilaremia is often associated with frequent symptoms such as Calabar swellings [4]. It would be beneficial for these populations to assess whether ALB, an inexpensive medication, could be used for curative purposes in patients with low microfilaremia as an alternative to IVM.

Therefore, the aim of this study was to evaluate the efficacy and safety of 400 mg dose of ALB compared to IVM for treating infected patients in Gabon.

## Material and methods

### Study sites and population

This survey was conducted among the rural populations of Woleu-Ntem, specifically in the departments of Ntem and Haut Ntem. Woleu-Ntem is a province located in the northern region of Gabon, a Central African country. The province has an equatorial climate characterized by hot, humid conditions and high rainfall. It is covered with old secondary forests and has rich and varied wildlife [19]. In Woleu-Ntem, the overall prevalence of *Loa loa* is 20.2%. The rural populations live along roads and rivers, with farming being their primary activity. Fishing and hunting are also common, which increases their exposure to Chrysops bites, the vectors responsible for transmitting *Loa loa* [20].

### Study design

This survey is a non-inferiority randomized control trial aimed at evaluating the efficacy and safety of a 400 mg dose of ALB taken daily for 30 days versus a single dose of IVM at 200μg/kg. The study was conducted from November 7, 2021, to April 1, 2022, in thirty-four villages within the province of Woleu-Ntem, located in northern Gabon. A preliminary visit was made to inform local authorities about the study’s purpose. After obtaining informed consent, participants’ sociodemographic and clinical data were collected. This included information on the study site, participant identification, age, sex, weight, height, and clinical symptoms related to loiasis, such as worm migration into the eye, Calabar swelling, adult worm migration under the skin, and pruritus.

#### Inclusion criteria

Eligible participants were individuals aged 18 to 75 years who were diagnosed with *Loa loa* infection confirmed by microscopy, with microfilarial counts ranging from ≥500 to ≤3500 mf/mL.

#### Exclusion criteria

Participants were excluded if they had acute systemic illnesses, suspected allergies to benzimidazoles, or were pregnant or nursing.

#### Blood collection and microfilaremic loiasis diagnosis

Venous blood was collected in an EDTA tube for parasitological diagnosis. A 10 µL blood sample was examined directly, and the leukoconcentration technique, as described by Ho Thi Sang and Petithory, was performed [21]. Microfilaremia was quantified as the number of microfilariae per milliliter of blood (mf/mL). Absence of microfilaremia was defined as the absence of detectable parasites following both microscopic techniques.

#### Eosinophil count

Eosinophils were counted from a thin blood smear prepared from a 5 µl sample. The count was expressed as a percentage based on 100 white blood cells observed under a light microscope. A count of ≥ 7% was considered indicative of hypereosinophilia.

### Randomization and treatment procedure

Individuals were randomly assigned to one of two treatment groups following stratification based on three levels of *Loa loa* microfilaremia: 500—1500; 1501—2500; 2501—3500 mf/ml, taking into account age, gender, and the level of microfilaremia. Treatment was administered as follows: patients either received a daily dose of one tablet of ALB (400 mg) for 30 days or a single calculated dose of IVM (200μg/kg) based on the patient’s weight. Both treatments were supplemented with 3 mg of antihistaminic-H2 daily for 7 days. Blood samples were collected before treatment initiation, and subsequently on days 2, 7, 14, and 30, to monitor microfilarial load and eosinophilia throughout the follow-up period.

### Monitoring of clinical signs and adverse events

Adverse events were systematically evaluated throughout the study and categorized by severity: mild if they caused minimal discomfort and did not disrupt daily activities, moderate if they interfered with daily activities to a noticeable extent, and severe if they significantly impaired the patient’s ability to carry out normal daily tasks.

### Efficacy assessments

Blood samples for direct examination (10 µL) and leukoconcentration were collected at baseline, prior to administration of the study drug, and on days 2, 7, 14, and 30 to assess microfilarial load. Given the diurnal periodicity of the filarial species, all efforts were made to collect blood samples between 10:00 am and 3:00 pm.

### Monitoring of eosinophil count

Eosinophils were counted from a thin blood smear using 5 µl of blood. Blood smears were conducted at baseline, prior to administration of the study drug, and on days 2, 7, 14, and 30 to assess the eosinophil levels.

### Ethical considerations

This study was conducted as part of the PHYLECOG project funded by the EDCTP2 program under reference TMA2019CDF-2730. The study protocol was reviewed and approved by the National Ethics Committee for Scientific Research. Written informed consent was obtained from each participant after explaining the study (Protocol No. 0053/2022/CNER/P/SG). The study is registered under ISRCTN14889921.

### Outcomes/Endpoints

The primary endpoint of this study was the reduction rate of microfilaremia. Non-inferiority between a monthly course of ALB and a single course of IVM was assessed based on the difference in reduction rates.

Treatment with ivermectin for microfilaremic loiasis typically results in rapid and sustained reduction, achieving a reduction rate of approximately 90% after 30 days, whereas placebo generally achieves a 50% reduction [22,23].

To establish non-inferiority, a margin of 20% was selected using the fixed margin method, which is half the difference in reduction rates observed between ivermectin and placebo.

The investigator monitored adverse events, categorizing them by type and severity. The primary safety endpoint was the occurrence of any adverse event following treatment initiation, regardless of its relationship to the study drug. The secondary safety endpoint involved tracking the evolution of eosinophil levels throughout the treatment period.

### Sample size

Based on an expected 73% reduction in microfilaremia with ALB and a 90% reduction with IVM [22–24], and assuming a first-species risk of 5.0% and a second-species risk of 10.0%, with a non-inferiority margin of 20%, it was determined that a minimum of 18 patients per group would be required for this study.

### Statistical analysis

Data processing was conducted using Microsoft Excel 2016, overseen by both an operator and a checker. R software version 4.3.0 was utilized for comprehensive data analysis.

Statistical analyses adhered to the principles outlined in the Consort 2010 statement, employing both per-protocol and intention-to-treat methodologies [25]. The per-protocol analysis focused exclusively on patients who completed their prescribed treatment regimen. Conversely, the intention-to-treat analysis included all randomized patients, irrespective of treatment completion.

Categorical data such as sex, symptoms, and adverse events were summarized as absolute values and percentages. These were compared using the proportion test, given the sample size was less than thirty, which is suitable for such analyses.

Continuous variables were evaluated for normality. Variables following a normal distribution, such as age, weight, height, BMI, and microfilaremia, were presented as mean ± standard deviation. Variables not following a normal distribution, like the eosinophil rate, were presented as median [1st percentile - 3rd percentile].

The reduction rate in microfilaremia was calculated as a percentage using the formula: ((Microfilaremia at D0 - Microfilaremia during follow-up) / Microfilaremia at D0) * 100.

Comparisons of microfilaremia or eosinophil rate between different time points were conducted using Student’s t-test for normally distributed variables or the Wilcoxon signed rank test for variables with non-normal distribution.

## Results

### Patients

Among the 1363 registered patients, 21 were excluded, leaving 1342 participants eligible for the study. Of these, 406 (30.2%) presented with *Loa loa* microfilaremia. Ultimately, 48 patients were enrolled, with 24 receiving ALB and 24 receiving IVM. However, during the follow-up period, 10 patients withdrew their consent (Fig 1).

**Fig. 1.**
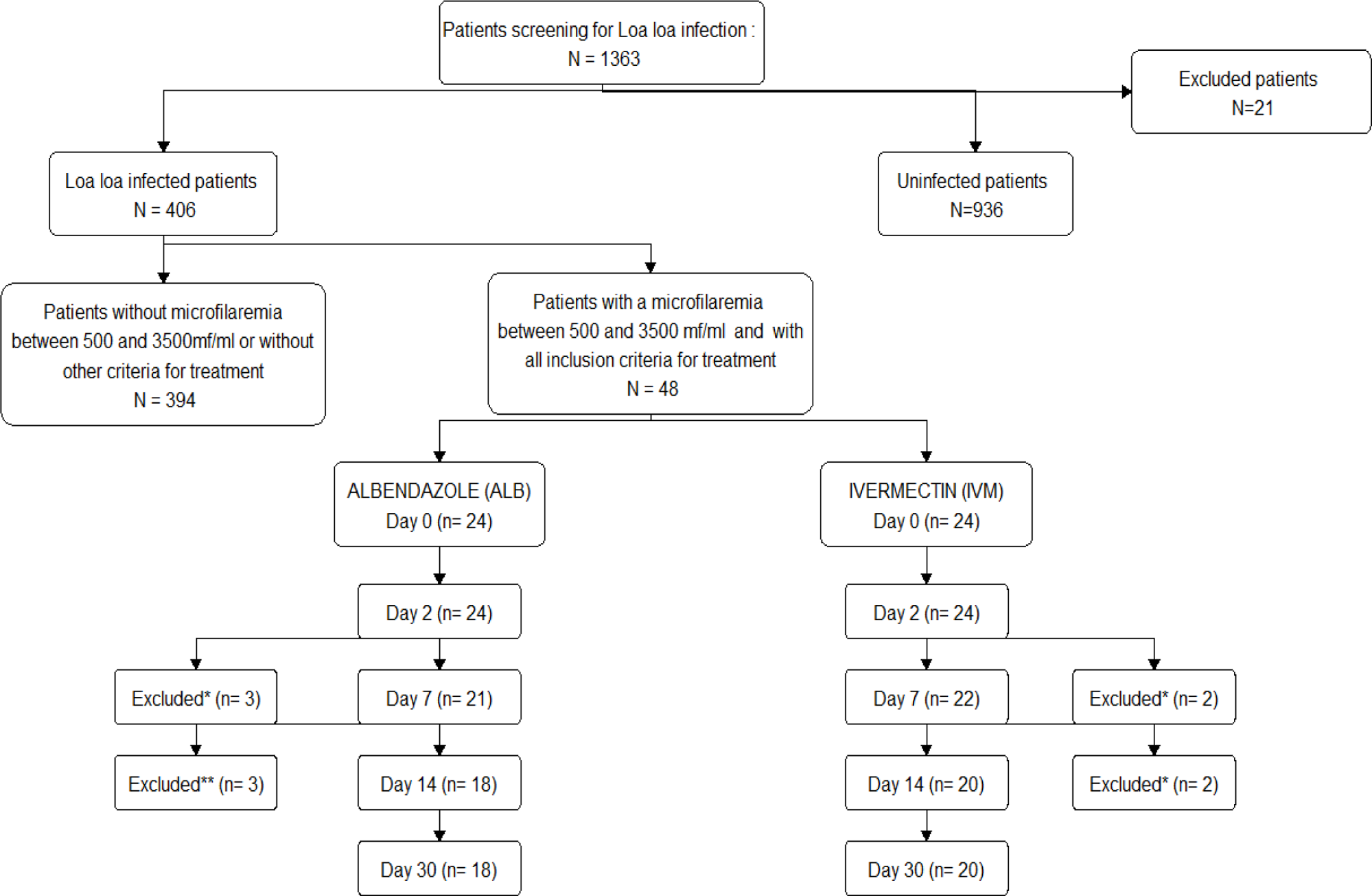
Study flowchart. * Refusal of venipuncture; **\*\*** Long-term traveler; **not visited**: could not be collected at the appropriate times for microfilaremia assessment, but present at the following visit.

### Demographic and baseline characteristics

The characteristics of the study population are summarized in Table 1. There were no significant differences between the two groups of patients (p > 0.05) in terms of sex, age, weight, height, microfilaremia, and eosinophil rate.

**Table 1.** Descriptive characteristics of the two treatment groups at the inclusion (D0)

|  | ALBENDAZOLE (n=22) | IVERMECTIN (n=24) | *p*-<br>value |
| --- | --- | --- | --- |
|  | n (%) | n (%) |  |
| Male | 17 (70.8) | 10 (41.7) | 0.08 |
| | Mean ± SD or<br>median [1<sup>st</sup> percentile - 3<sup>rd</sup> percentile] | Mean ± SD or<br>median [1<sup>st</sup> percentile - 3<sup>rd</sup> percentile] | |
| Age (years) | 49.8±12.5 | 53.0±13.6 | 0.4 |
| Weight (Kg) | 64.3±10.3 | 64.5±10.7 | 0.9 |
| Height (m) | 1.6±0.1 | 1.6±0.1 | 0.2 |
| BMI (Kg/m<sup>2</sup>) | 23.6±3.2 | 24.6±4.2 | 0.4 |
| Microfilaremia (mf/mL) | 1150 [700 - 1625] | 1350 [875 - 2000] | 0.5 |
| Eosinophil rate (%) | 16.0 [10.5 - 21.0] | 16.0 [12.5 - 25.5] | 0.5 |

SD: standard deviation

### Per-protocol analysis

#### Efficacy of treatments on the reduction of microfilaremia

Both treatment groups showed significant reductions in mean microfilaremia from day 0 to day 30. Specifically, the ALB group exhibited a reduction of 82.3% (1339 to 237 mf/ml), while the IVM group showed a reduction of 90.4% (1395 to 134 mf/ml) (p < 0.001) (Fig 2.a). The risk difference between the two treatments was 8.1% [95% CI: 16.8; −0.6%], and the 95% confidence interval did not exceed the margin of non-inferiority.

**Fig 2.a.**
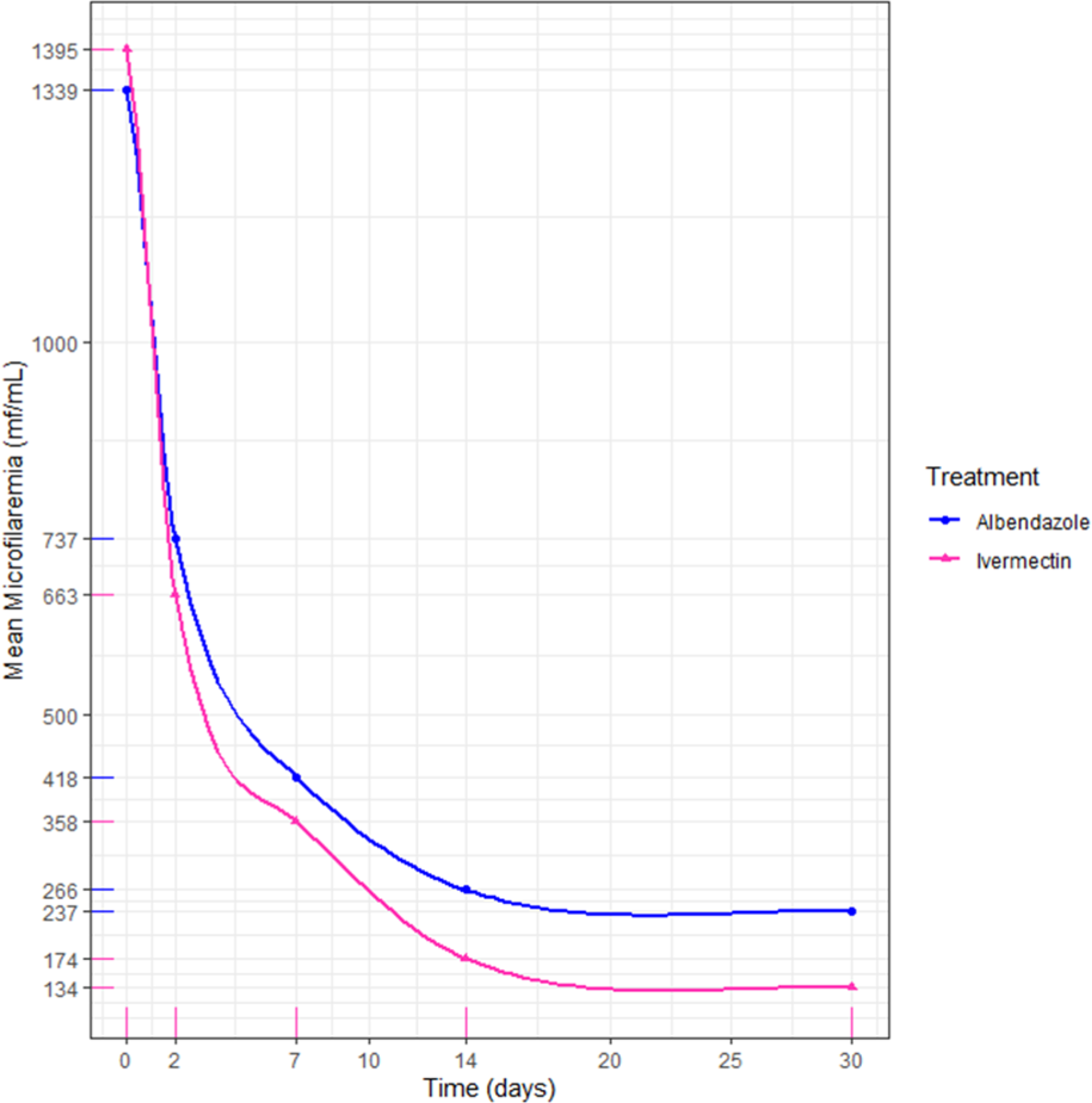
Comparison of the albendazole and ivermectin parasitological cure rate (Per-protocol). The blue line on the chart represents the microfilaremia levels of patients treated with albendazole, while the pink line represents those treated with ivermectin.

**Fig 2.b.**
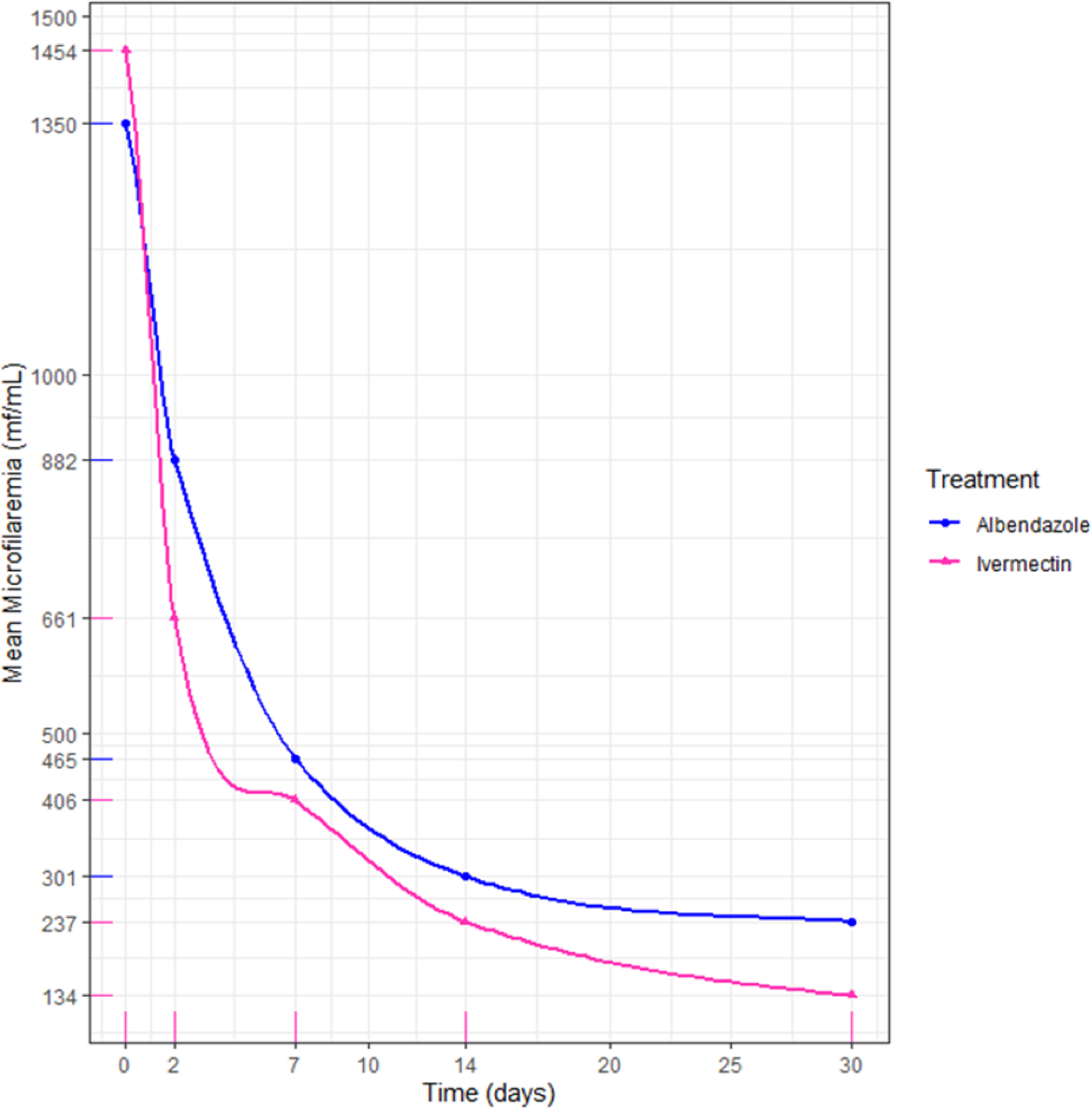
Comparison of the albendazole and ivermectin parasitological cure rate (Intention-to-treat). In this line chart, the blue line represents the microfilaremia levels of patients treated with albendazole, while the pink line represents those treated with ivermectin.

### Intention-to-treat analysis and secondary outcomes

#### Efficacy of treatments on the reduction of microfilaremia

The mean microfilaremia in patients from both treatment groups decreased significantly by 82.4% (1350 to 237 mf/ml) and 90.8% (1454 to 134 mf/ml) from day 0 to day 30 in the ALB and IVM groups, respectively (p < 0.001) (Fig 2.b). The risk difference between the two treatments was 8.4% [95% CI: 16.5; 0.7%], and the 95% confidence interval did not exceed the margin of non-inferiority.

#### Incidence and progression of adverse events and symptoms during treatment

At baseline, approximately half of the patients in both groups reported symptoms: 41.7% (n=10) in the ALB group and 54.2% (n=13) in the IVM group (p=0.6). The most frequent symptom was pruritus, noted in 90.0% (n=9/10) of patients in the ALB group and 69.2% (n=9/13) in the IVM group. Over the course of follow-up, the number of patients experiencing symptoms decreased, with no patients reporting symptoms by day 30 in either group (Fig 3.a).

**Fig 3.a.**
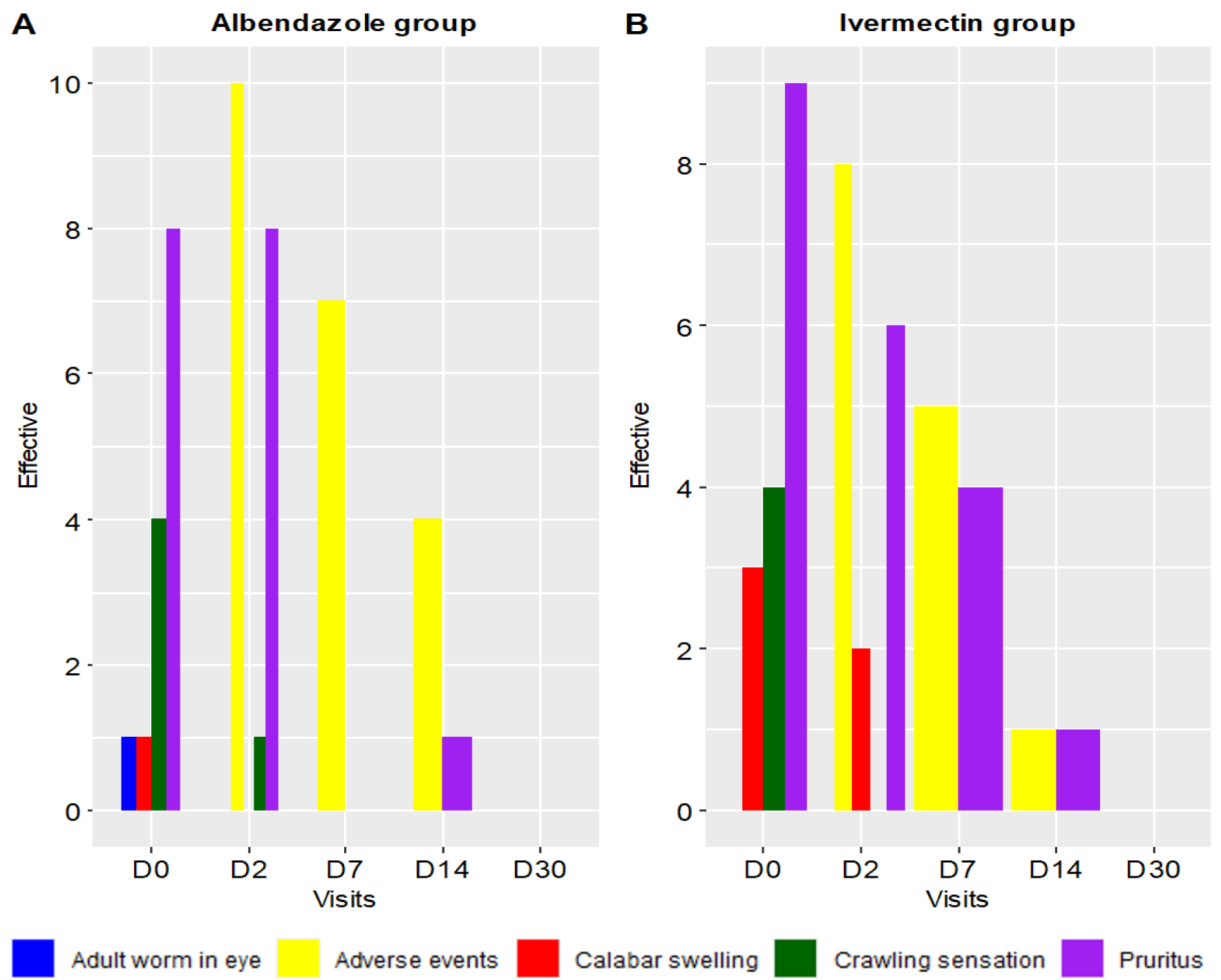
Changes in clinical signs and adverse events over the 30-day follow-up period. The histogram shows adult worms in the eye with blue bars, adverse events with yellow bars, Calabar swelling with red bars, crawling sensation with green bars, and pruritus with purple bars.

In terms of adverse events, a total of 10 patients (41.7%) in the ALB group and 8 patients (33.3%) in the IVM group reported adverse events by day 2 (p=0.8) (Fig 3.b). The most common adverse event in the ALB group was asthenia, reported by 80.0% (n=8/10) of those experiencing adverse events, followed by increased appetite in 20.0% (n=2/10). In the IVM group, the most common adverse events were asthenia, reported by 50.0% (n=4/8), and headache, reported by 25.0% (n=2/8) (Fig 3.b).

**Fig 3.b.**
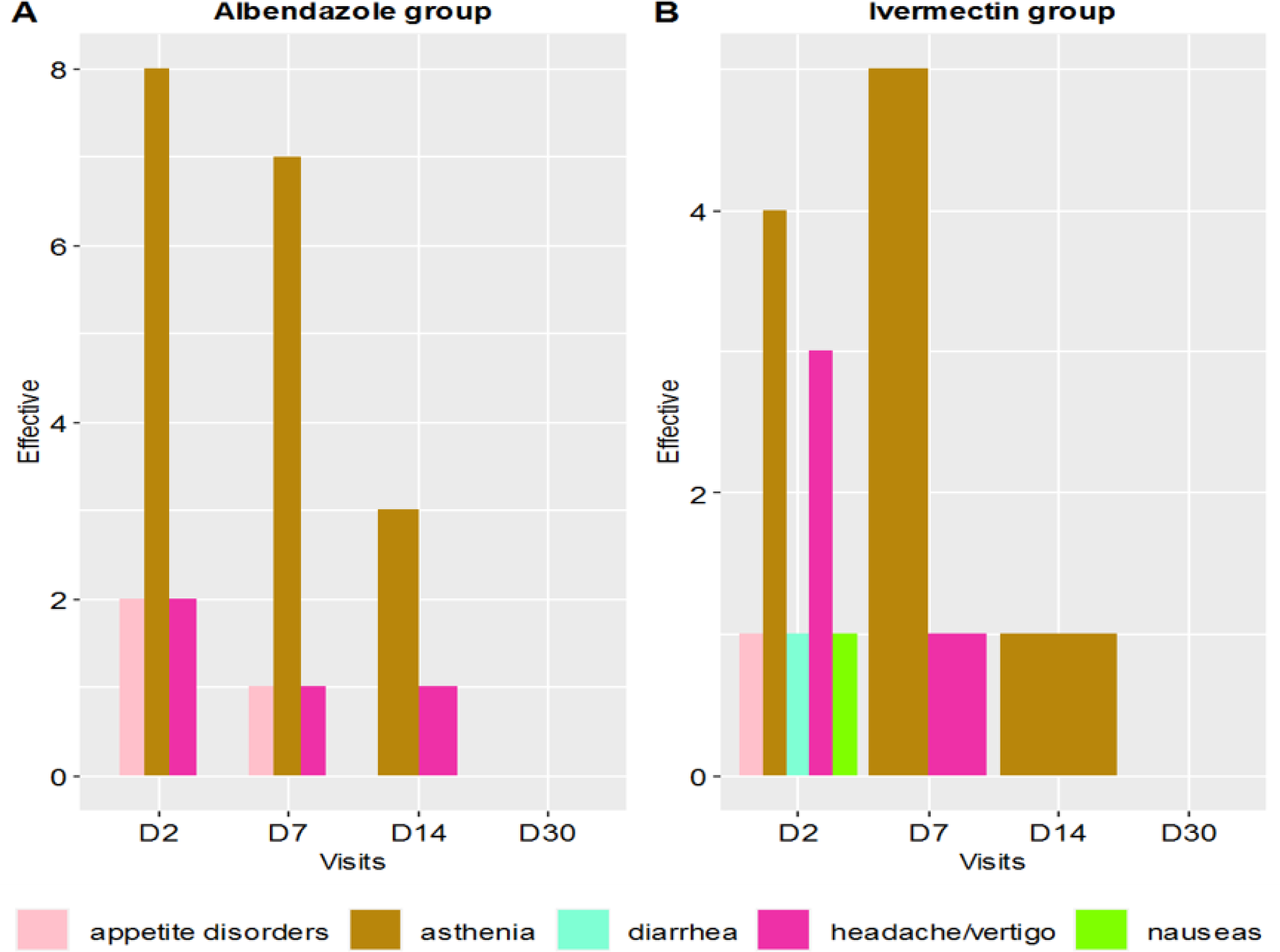
Changes in clinical adverse events over the 30-day follow-up period. The histogram shows appetite disorders with pink bars, asthenia with dark gold bars, diarrhea with an aquamarine bar, headache and/or vertigo with maroon bars, and nausea with a chartreuse green bar.

#### Efficacy of treatments on eosinophil level reduction

In both groups, eosinophil rates decreased by Day 30, although the difference was not statistically significant (p > 0.05). In the albendazole group, eosinophil rates initially increased from Day 0 to Day 2, followed by a decline across subsequent visits, reducing from a median rate of 16% to 7% (Fig 8). Conversely, in the ivermectin group, eosinophil rates remained stable from Day 0 to Day 14, with a decrease observed by Day 30 from a median rate of 16% to 4% (Fig 4).

**Fig 4.**
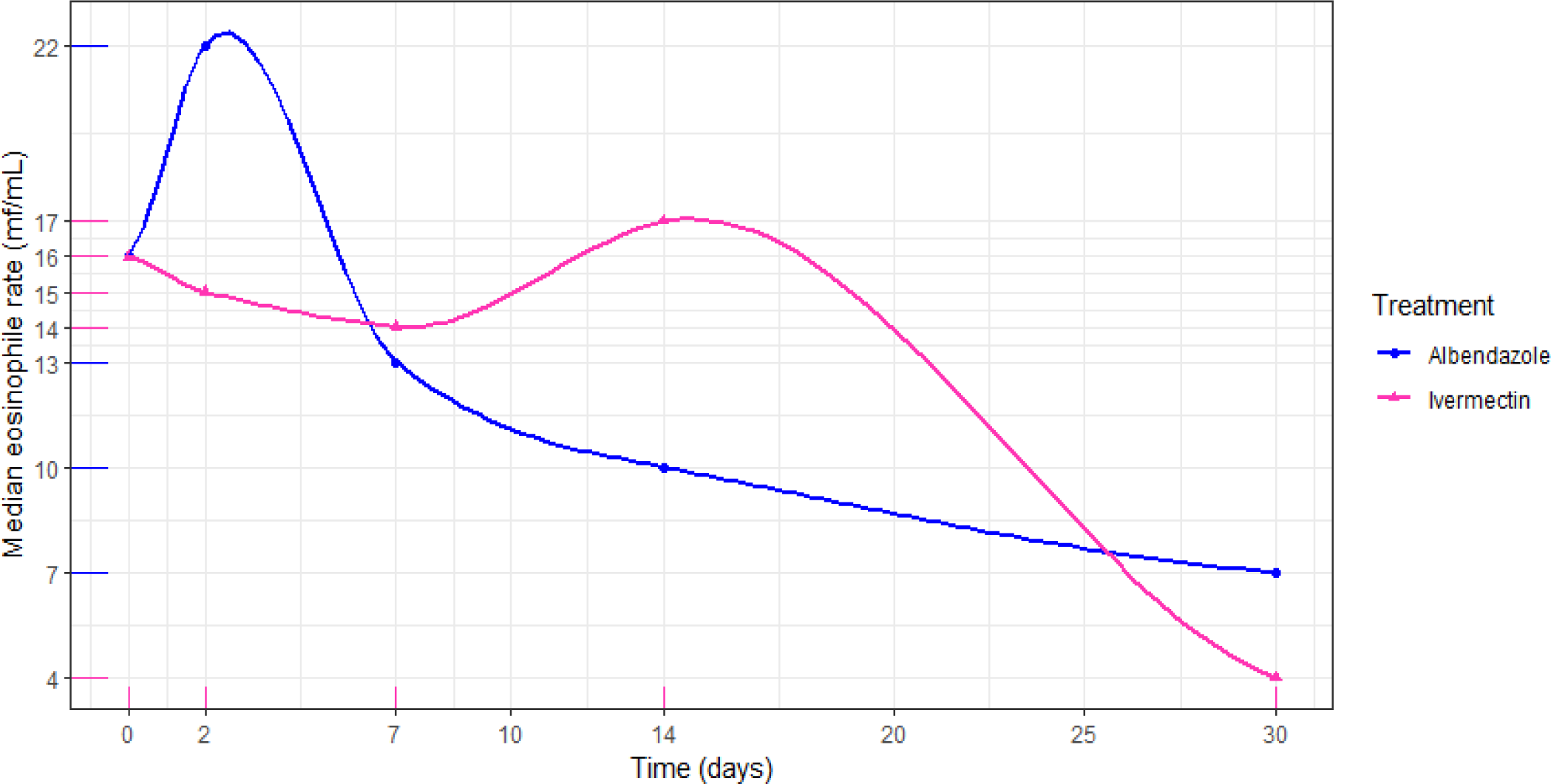
Eosinophil rate progression by treatment. The blue line on the chart depicts the eosinophil rate of patients treated with albendazole, while the pink line represents those treated with ivermectin.

## Discussion

This clinical trial represents the first comparison of a 30-day treatment regimen with ALB and IVM. The primary objective was to evaluate ALB’s efficacy and safety as a potential non-inferior alternative to IVM. This is particularly relevant for landlocked rural populations who may have easier access to ALB than IVM.

In the intention-to-treat analysis of this trial, patients in both the ALB and IVM groups experienced a significant decrease in mean microfilaremia from day 0 to day 30. The reduction was more pronounced among those treated with IVM, with a risk difference of 8.6%. However, the 95% confidence interval for this difference did not reach or exceed the non-inferiority margin of 20%. These results confirm the non-inferiority of monthly ALB treatment compared to IVM.

For the per-protocol analysis, the same trend was observed. Patients’ mean microfilaremia significantly decreased from day 0 to day 30 in both the ALB and IVM groups. The reduction was more pronounced in patients treated with IVM, with a risk difference of 8.1%. However, the 95% confidence interval for this risk difference did not exceed the non-inferiority margin of 20%, confirming the non-inferiority of monthly ALB treatment compared to IVM.

During the follow-up, the number of patients with symptoms decreased, and no patient exhibited any loiasis symptoms by day 30 in either group. This highlights an adequate clinical response to both treatments. However, these findings contrast with the results of the retrospective observational study by Gobbi et al., where 31.3% of patients treated with a single dose of IVM and 50% of those treated with ALB (400 mg/day) were still symptomatic after 30 days of treatment [26].

Less than half of the patients in both treatment groups experienced adverse events during the follow-up period, with asthenia being the most common in both groups. The frequency of these adverse events decreased over subsequent visits, indicating the treatments’ safety, all of which were mild. Similar findings have been reported in prior studies using comparable dosages, where no participants experienced serious adverse events [26,27].

Regarding the impact of both treatments on eosinophil counts, there was no statistically significant reduction observed in either treatment group during the follow-up period. These results are consistent with Herrick et al., who found no decrease in eosinophil rates after two weeks of IVM treatment [28]. They also align with Klion et al., who reported no reduction in eosinophil rates after 21 days of ALB treatment [24]. However, despite the lack of significant decrease over the 30-day treatment period, there appeared to be a negative correlation between treatment duration and eosinophil rates.

These findings suggest that monthly treatment with ALB could serve as a viable alternative for rural populations lacking easy access to IVM and limited financial resources. The study’s strengths include its randomized design, prospective registration, and comprehensive analysis employing both per-protocol and intention-to-treat methods. However, the study also had its limitations. Firstly, due to the nature of the treatments, blinding was not feasible. Additionally, being a non-inferiority study focused on comparing microfilaremia reduction rates between treatments, it had inherent constraints in assessing other potential clinical endpoints.

## Conclusion

The prevalence of *Loa loa* infection is notably high in the investigated villages of the Woleu-Ntem province, Gabon. While IVM treatment exhibited a more pronounced microfilaricidal effect, the difference in risk between the two treatments, as indicated by the associated 95% confidence interval, did not exceed the margin of inferiority. This confirms the non-inferiority of ALB treatment. Moreover, ALB treatment demonstrated an adequate clinical response and exhibited good safety profiles similar to IVM.

## Data Availability

All data produced in the present study are available upon reasonable request to the authors

## Acknowledgments

The authors extend their sincere gratitude to all the staff at the Centre de Recherche biomédicale En pathogènes Infectieux et Pathologies Associées (CREIPA) and the Unité Mixte de Recherche sur les Agents Infectieux et leur Pathologie (UMRAIP) for their invaluable support in participant recruitment in Bitam, and for their assistance with sample analysis in both Libreville and Bitam. Special thanks are also due to the management of the Bifolossi Medical Center for generously hosting our study. We extend our appreciation to the village chiefs and other relevant authorities for their unwavering support throughout this research. Finally, heartfelt thanks to all the participants whose cooperation made this study possible.

## Author contributions

**Conceptualization:** Denise Patricia Mawili-Mboumba, Marielle Karine Bouyou Akotet, Noé Patrick M’Bondoukwé

**Data curation:** Luccheri Ndong Akomezoghe, Noé Patrick M’Bondoukwé

**Formal analysis:** Luccheri Ndong Akomezoghe, Noé Patrick M’Bondoukwé

**Funding acquisition:** Marielle Karine Bouyou Akotet, Noé Patrick M’Bondoukwé

**Investigation:** Luccheri Ndong Akomezoghe, Mérédith Flore Ada Mengome, Roger Hadry Sibi Matotou, Bridy Chesly Moutombi Ditombi, Valentin Migueba, Jacques Mari Ndong Ngomo, Noé Patrick M’Bondoukwé

**Methodology:** Luccheri Ndong Akomezoghe, Denise Patricia Mawili-Mboumba, Noé Patrick M’Bondoukwé

**Project administration:** Denise Patricia Mawili-Mboumba, Marielle Karine Bouyou Akotet

**Resources:** Marielle Karine Bouyou Akotet

**Supervision:** Denise Patricia Mawili-Mboumba, Marielle Karine Bouyou Akotet

**Validation:** Denise Patricia Mawili-Mboumba, Marielle Karine Bouyou Akotet

**Visualization:** Luccheri Ndong Akomezoghe

**Writing-original draft:** Luccheri Ndong Akomezoghe, Noé Patrick M’Bondoukwé, Denise Patricia Mawili-Mboumba

**Writing-review and editing:** Luccheri Ndong Akomezoghe, Marielle Karine Bouyou Akotet, Denise Patricia Mawili-Mboumba, Noé Patrick M’Bondoukwé

